# Advanced Cardiac Imaging in Treatment Decision-Making for Severe Left Ventricular Dysfunction: A Systematic Review and Meta-Analysis

**DOI:** 10.1101/2025.05.12.25327464

**Authors:** Tabitha Kusi-Yeboah, Dalton J. Bally, Milindu Wickramarachchi, Michael Chiu, Kieran Williams, Sebastian Möbus, Anakha Varma, Amogh Bandla, Maria Taiwo, Hasan Butt, Arnav Sharma

## Abstract

This review explores whether assessing myocardial viability before coronary revascularization meaningfully impacts outcomes in patients with ischemic cardiomyopathy. By synthesizing evidence from studies employing imaging techniques such as CMR, PET, SPECT, and dobutamine stress echocardiography, we examined associations between viability status and improvements in cardiac function and survival. Our findings suggest that patients with viable myocardium tend to derive greater benefit from revascularization, particularly in terms of left ventricular recovery and reduced mortality. These results highlight the potential value of viability imaging in guiding treatment decisions and support its selective use in clinical practice.

## Introduction And Background

Ischemic cardiomyopathy (ICM), characterized by impaired left ventricular (LV) function due to chronic coronary artery disease, remains a leading contributor to heart failure and cardiovascular mortality worldwide [1]. Effective management hinges critically on accurately identifying patients likely to benefit from revascularization, which involves distinguishing viable myocardium capable of functional recovery from irreversibly scarred tissue [2–4]. Traditional methods such as echocardiography, although widely used, possess limited spatial resolution and insufficient myocardial tissue characterization, constraining their diagnostic precision in this clinical scenario [5].

Advanced cardiac imaging modalities-particularly cardiac magnetic resonance (CMR) and positron emission tomography (PET)-have emerged as indispensable tools [6]. for improving therapeutic decisions by providing detailed insights into myocardial viability [7,8]. CMR, utilizing late gadolinium enhancement (LGE), effectively differentiates viable myocardium from fibrotic segments and quantifies scar burden, thereby guiding appropriate patient selection for revascularization [9,10]. PET imaging complements anatomical detail with metabolic data [11], specifically via 18F-fluorodeoxyglucose (FDG), identifying hibernating myocardium that retains metabolic activity despite contractile dysfunction and thus is potentially salvageable with revascularization [3,4].

Despite robust evidence supporting their diagnostic utility [12], the extent to which advanced viability imaging influences clinical outcomes such as functional recovery and survival remains incompletely defined. While prior observational studies have suggested improved survival associated with imaging-guided revascularization, recent randomized evidence, such as the STICH trial, has cast doubt on survival benefits from coronary artery bypass grafting (CABG) in patients with viable myocardium [8,10]. This evolving evidence base underscores the complexity of imaging-informed clinical decision-making and the ongoing need to clarify the role of advanced imaging modalities.

Accordingly, this systematic review and meta-analysis aim to address critical gaps in our understanding by evaluating the diagnostic and prognostic utility of CMR and PET in patients with ICM undergoing revascularization. Specifically, we investigate the ability of these modalities to predict left ventricular ejection fraction (LVEF) improvement and all-cause mortality, comparing imaging-guided revascularization strategies against non-guided approaches. By synthesizing current literature, this review seeks to inform an evidence-based framework for effectively integrating viability imaging into individualized treatment decisions for severe LV dysfunction.

## Review

### Methods

#### Eligibility Criteria

This systematic review included studies enrolling adult patients (aged ≥18 years) diagnosed with ischemic cardiomyopathy (ICM) and severely reduced left ventricular ejection fraction (LVEF <35%) who underwent coronary revascularization, either via coronary artery bypass grafting (CABG) or percutaneous coronary intervention (PCI). Eligible studies involved advanced cardiac imaging modalities, such as cardiac magnetic resonance imaging (CMR) with late gadolinium enhancement (LGE), positron emission tomography (PET) utilizing 18F-fluorodeoxyglucose (FDG), single-photon emission computed tomography (SPECT), and dobutamine stress echocardiography, which were employed to assess myocardial viability prior to revascularization.

To be included in this review, studies had to evaluate the association between imaging-guided revascularization and one of the two primary outcomes: (1) improvement in LVEF following revascularization, or (2) all-cause mortality. Both randomized controlled trials (RCTs) and prospective or retrospective cohort studies were eligible for inclusion. Only studies published in English between January 2013 and March 2025 were considered.

Studies were excluded if they: (1) focused on non-ischemic cardiomyopathy; (2) enrolled patients with preserved LVEF (>35%); (3) used imaging without influencing the treatment strategy; (4) lacked extractable outcome data, such as hazard ratios (HRs), odds ratios (ORs), or mean differences in LVEF; or (5) did not stratify by viable versus non-viable myocardium. Case reports, editorials, narrative reviews, conference abstracts without full text, and studies that did not assess viability-based imaging-guided revascularization were also excluded.

#### Search Strategy

A comprehensive systematic search was conducted across PubMed (2013-2025) and Embase (1974-2025) to identify peer-reviewed studies relevant to the review. Search terms were formulated in accordance with the PICO framework, combining keywords for the population (e.g., “ischemic cardiomyopathy,” “heart failure,” “reduced ejection fraction”), intervention (e.g., “cardiac MRI,” “late gadolinium enhancement,” “FDG-PET,” “viability imaging”), and outcomes (e.g., “mortality,” “LVEF,” “functional recovery,” “revascularization,” “survival”). Filters were applied to restrict results to clinical trials and cohort studies. A detailed search strategy can be found in the Supplementary Appendix.

#### Study Selection and Data Extraction

After removing duplicate entries, titles and abstracts were initially screened by a single reviewer (Tabitha Kusi-Yeboah). Full-text articles of potentially eligible studies were independently assessed by two pairs of reviewers (Dalton Bally/Milindu Wickramarachchi, Kieran Williams/Anakha Varma, Hasan Butt/Amogh Bandla, Maria Taiwo/Michael Chiu, Sebastian Möbus/Arnav Sharma) based on pre-specified eligibility criteria (Figure *1*). Any disagreements were resolved through discussion or by a third reviewer (Tabitha Kusi-Yeboah). Data were extracted using a standardized form, capturing key study characteristics, including author, publication year, country of study, design, population demographics, imaging modality, intervention and control definitions, and duration of follow-up.

**FIGURE 1:**
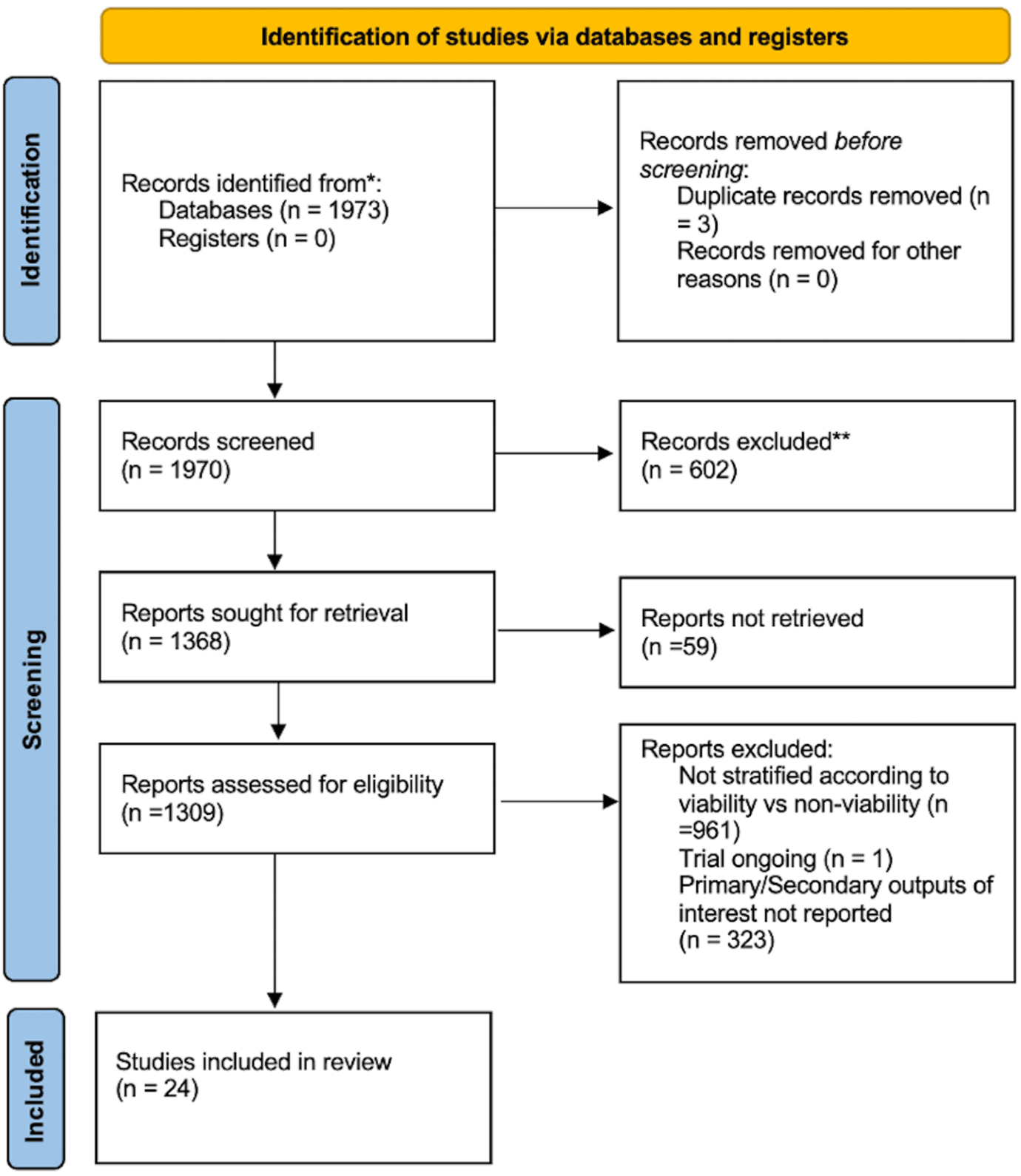
PRISMA 2020 Flow Diagram for Study Selection. Flow diagram illustrating the process of study identification, screening, eligibility assessment, and final inclusion in the systematic review. A total of 1,973 records were identified through database searches. After removing duplicates and exclusions, 24 studies met eligibility criteria and were included in the final synthesis. Adapted from Page MJ, et al. BMJ. 2021;372:n71. doi: 10.1136/bmj.n71

The primary outcomes of interest were the absolute change in LVEF following imaging-guided revascularization and all-cause mortality. For studies reporting these outcomes, effect estimates were recorded as mean differences (MDs), hazard ratios (HRs), or odds ratios (ORs), along with 95% confidence intervals (CIs). If effect estimates were not directly reported, values were derived from raw event counts or Kaplan-Meier curves using established methods, including log transformation of odds ratios or relative risks to estimate HRs [13].

#### Outcomes of Interest

The primary outcomes included: (1) Improvement in LVEF following imaging-guided revascularization, and (2) All-cause mortality, comparing imaging-guided versus non-guided revascularization strategies.

Secondary outcomes of interest included a range of clinical and functional measures. These included the comparative effectiveness of CMR versus PET in predicting post-revascularization LVEF improvement. Additionally, we examined exercise capacity as measured by METs and impact of revascularisation on perfusion update.

#### Risk of Bias Assessment

The methodological quality of included studies was independently assessed by two reviewers. Randomized controlled trials were evaluated using the Cochrane Risk of Bias 2.0 (RoB 2.0) tool ([14] which appraises five domains: the randomization process, deviations from intended interventions, missing outcome data, measurement of the outcome, and selection of the reported result. Observational cohort studies were assessed using the Risk Of Bias In Non-randomized Studies - of Interventions (ROBINS-I) tool ([15]. This tool evaluates bias across seven domains: confounding, participant selection, classification of interventions, deviations from intended interventions, missing data, outcome measurement, and selection of the reported result. Disagreements were resolved through discussion and consensus.

#### Statistical Analysis

Meta-analysis was conducted using Review Manager (RevMan) version 5.4. For continuous outcomes (e.g., LVEF change), pooled effect sizes were expressed as mean differences (MDs) with corresponding 95% confidence intervals. For binary outcomes (e.g., mortality), hazard ratios (HRs) were extracted directly or calculated from raw event data. Where HRs were unavailable, odds ratios (ORs) were derived and approximated as HRs for meta-analysis.

A random-effects model was employed for all pooled analyses using the Restricted Maximum Likelihood (REML) estimator to account for heterogeneity across studies [16]. This method was chosen due to its accuracy in estimating between-study variance (τ²), particularly when a limited number of studies are included. Statistical heterogeneity was quantified using the I² statistic, and Cochran’s Q test (Chi²) was reported. A p-value <0.10 or I² >50% was considered indicative of substantial heterogeneity.

#### Subgroup and Sensitivity Analyses

Prespecified subgroup analyses included comparisons by imaging modality. However, due to the limited number of studies in certain subgroups, formal tests for subgroup differences were not performed. Sensitivity analyses were performed by sequentially excluding individual studies to evaluate the robustness of the pooled estimate. These analyses confirmed the stability and statistical significance of the results.

## Results

### Study Selection and Characteristics

A total of 24 studies were included in this systematic review, of which 22 contributed to the quantitative meta-analysis. Thirteen studies reported changes in left ventricular ejection fraction (LVEF) [17–29], eight studies provided data on regional wall motion score (RWMS) [18,20–22,24–26,30] and nine studies assessed all-cause mortality [31–38]. Across all studies, advanced imaging modalities were employed to assess myocardial viability prior to revascularization, including cardiac magnetic resonance imaging (CMR), positron emission tomography (PET), single-photon emission computed tomography (SPECT), and dobutamine stress echocardiography. The included patient populations varied in size and baseline characteristics, but all contributed to the pooled or qualitative evaluation of functional and prognostic outcomes based on myocardial viability status.

### LVEF Change

The overall pooled analysis of LVEF change following revascularization revealed a statistically significant improvement in the viable myocardium group compared to the non-viable group. The mean difference in LVEF was 4.03% (95% CI: 3.76-4.31), with substantial heterogeneity across studies (I² = 98%, p < 0.00001) (Figure *2*). This suggests that the presence of viable myocardium is associated with a greater improvement in LVEF after revascularization, reinforcing the utility of viability imaging in guiding revascularization decisions.

**FIGURE 2:**
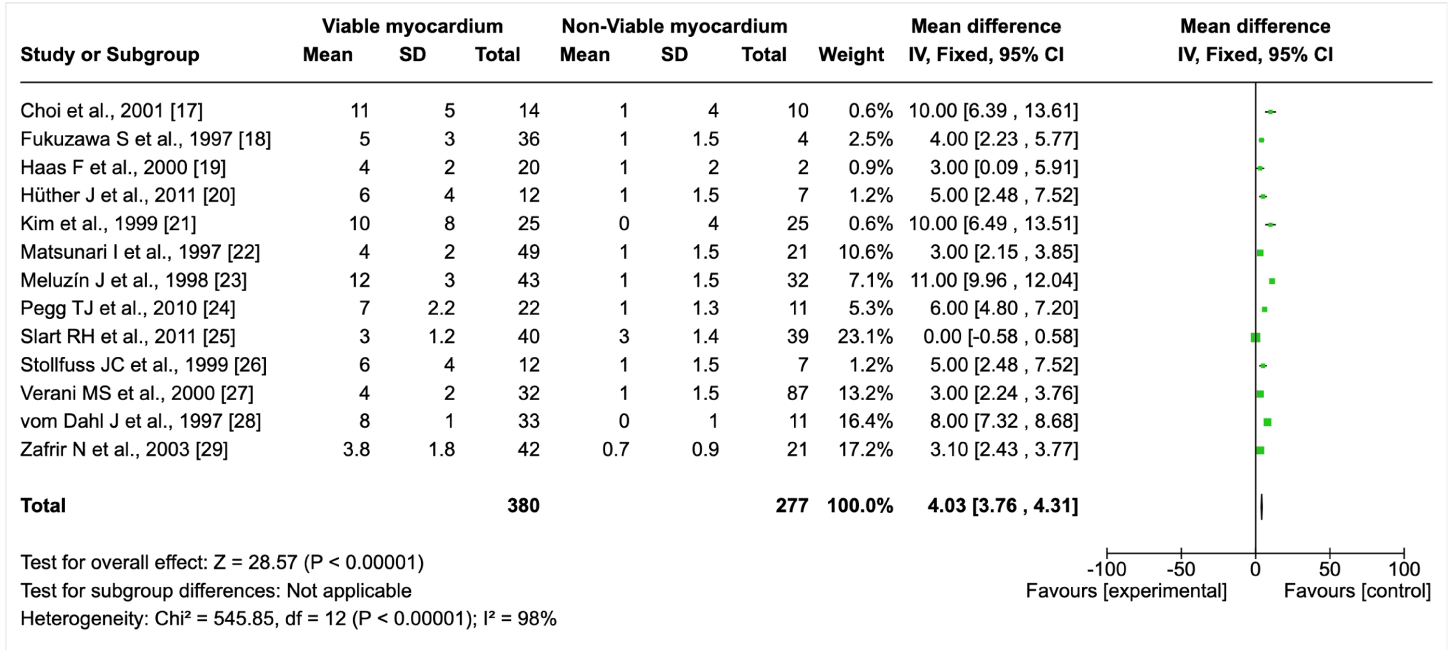
Forest Plot of Mean Change in Left Ventricular Ejection Fraction (LVEF) Following Revascularization in Patients With Viable vs. Non-Viable Myocardium. The forest plot summarizes pooled mean differences in LVEF improvement across 13 studies comparing patients with viable myocardium to those with non-viable myocardium. The overall effect favors the viable group, with a statistically significant mean difference of 4.03% (95% CI: 3.76–4.31). Fixed-effects model used; heterogeneity was substantial (I² = 98%). Abbreviations: SD – standard deviation; CI – confidence interval; IV – inverse variance.

### Subgroup Analysis by Imaging Modality

A detailed subgroup analysis was conducted to explore the impact of different imaging modalities on the LVEF improvement between viable and non-viable myocardium. All modalities showed significantly greater LVEF improvement in the viable group (Figure *3*)

**FIGURE 3:**
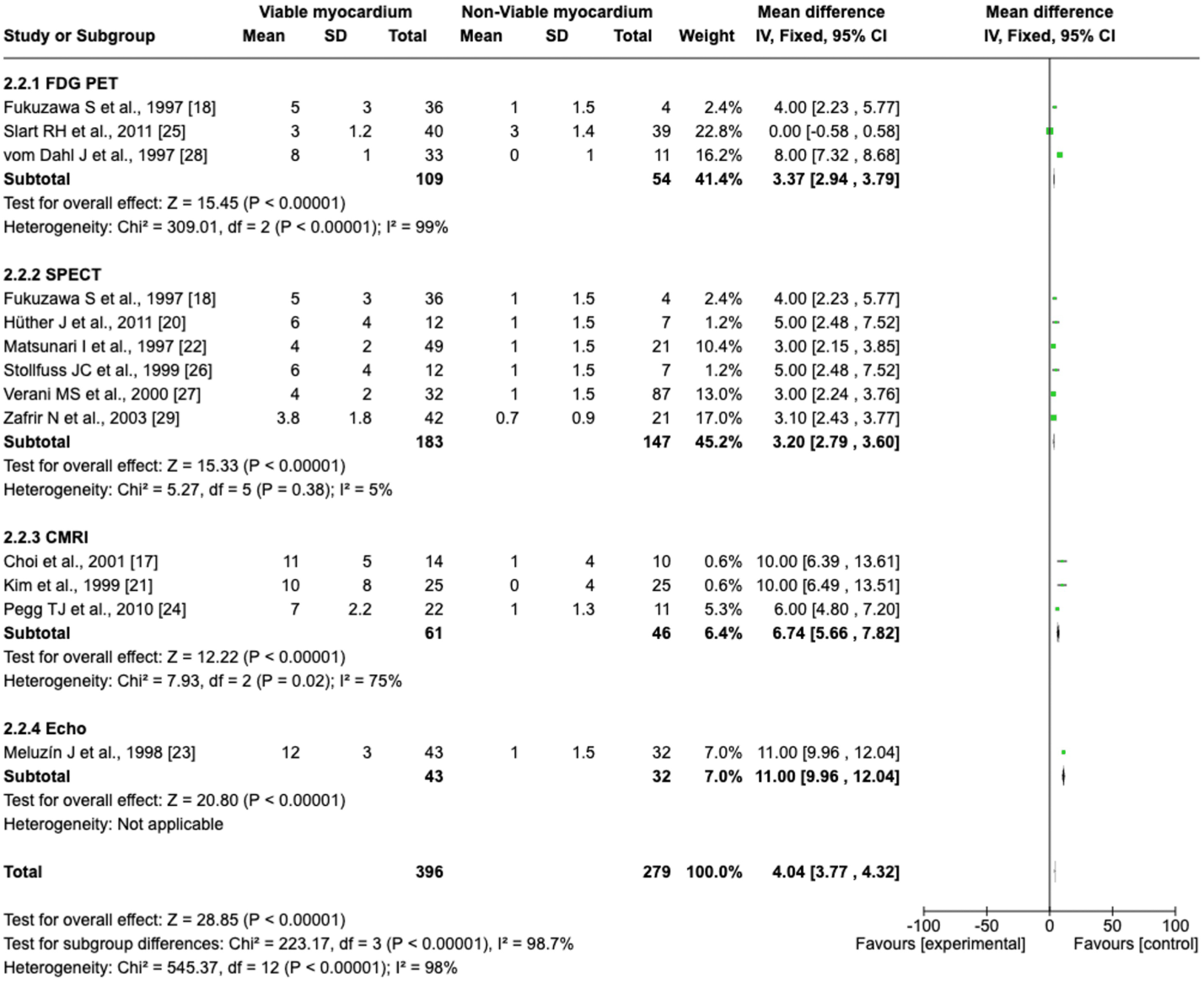
Subgroup Analysis of LVEF Improvement by Imaging Modality. Forest plot illustrating mean differences in left ventricular ejection fraction (LVEF) following revascularization among patients with viable versus non-viable myocardium, stratified by imaging modality. All modalities (FDG-PET, SPECT, cardiac magnetic resonance imaging [CMRI], and dobutamine stress echocardiography) showed significantly greater LVEF improvement in the viable group. The greatest effect was observed with echocardiography (mean difference: 11.00%, 95% CI: 9.96–12.04), followed by CMRI (6.74%, 95% CI: 5.66– 7.82). Between-subgroup heterogeneity was statistically significant (p < 0.00001). Abbreviations: SD – standard deviation; CI – confidence interval; IV – inverse variance.

FDG PET: The subgroup of studies utilizing FDG-PET showed a significant difference in LVEF improvement between the viable and non-viable groups, with a mean difference of 3.37% (95% CI: 2.94-3.79). The heterogeneity within this group was very high (I² = 99%, p < 0.00001), indicating variation across studies.

SPECT: Studies using SPECT also demonstrated a significant improvement in LVEF in the viable myocardium group. The mean difference for this modality was 3.20% (95% CI: 2.79-3.60), with substantial heterogeneity (I² = 55%, p < 0.00001).

CMR: The CMR subgroup analysis revealed a mean difference of 6.74% (95% CI: 5.66-7.82) in LVEF between viable and non-viable myocardium. The studies in this subgroup had moderate heterogeneity (I² = 75%, p = 0.02).

Echo: The subgroup of studies using dobutamine stress echocardiography showed a mean difference of 11.00% (95% CI: 9.96-12.04), suggesting a strong improvement in LVEF in the viable myocardium group compared to the non-viable group. This result was consistent across the included studies, with no significant heterogeneity.

### Overall Comparison by Imaging Modality

The pooled analysis from all imaging modalities revealed a consistent finding that imaging-guided revascularization, particularly when viability is identified, leads to a greater improvement in LVEF compared to non-viable myocardium. The mean difference in LVEF across all imaging modalities was 4.04% (95% CI: 3.77-4.32), reinforcing the role of viability imaging in guiding revascularization.

### Sensitivity Analysis - LVEF

Sensitivity analyses were conducted to assess the robustness of the observed LVEF benefit under a fixed-effects model. Across all permutations of study exclusion, the effect of viability on post-revascularization LVEF remained statistically significant, with mean differences ranging from 3.25% (95% CI: 2.95-3.56) to 5.25% (95% CI: 4.93-5.56). Notably, exclusion of Slart et al., [27], the study with the largest weight and a null result, resulted in the highest observed effect size (5.25%), indicating its substantial influence on the pooled estimate. However, even with its inclusion, the effect remained significant and directionally consistent. This underscores the robustness of the association between viable myocardium and improved LVEF, regardless of individual study influence. Heterogeneity persisted across models (I² = 96-98%), likely reflecting methodological and population-level variability across included studies.

### RWMS Overall Analysis

In addition to assessing LVEF, a pooled analysis was performed for changes in regional wall motion score (RWMS) following revascularization. This included 282 patients with viable myocardium and 187 with non-viable myocardium. The overall pooled mean difference in RWMS between the viable and non-viable groups was -0.05 (95% CI: -0.45 to 0.35), indicating no statistically significant difference (p = 0.80) [17–29]. Heterogeneity was substantial (I² = 100%, Tau² = 0.34), driven largely by marked variability in study outcomes and effect sizes (Figure *4*). Notably, the study by Murashita et al., [30] again emerged as a clear outlier, reporting an exaggerated RWMS improvement (mean difference = 2.40 [95% CI: 1.32 to 3.48]). This outlier substantially influenced the overall pooled estimate and contributed disproportionately to between-study variance. The findings suggest that, while viable myocardium may confer regional functional recovery, the presence of extreme variability and influential studies can obscure the underlying association in aggregate-level analysis.

**FIGURE 4:**
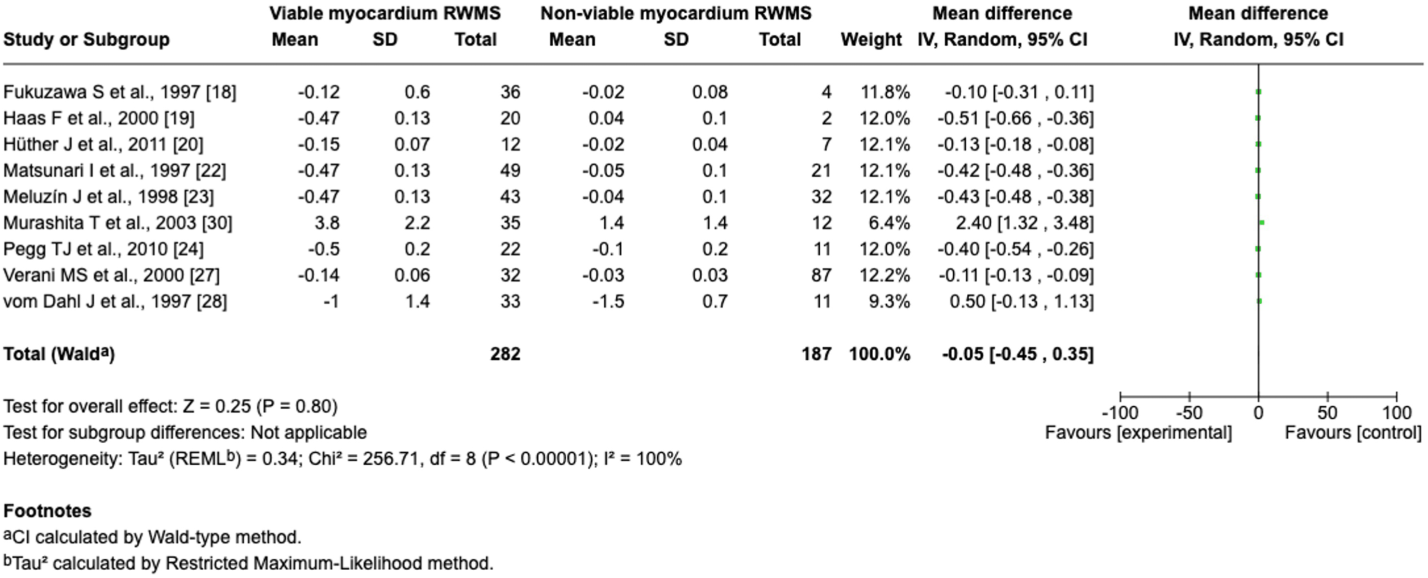
Forest Plot of Regional Wall Motion Score (RWMS) Change After Revascularization in Patients With Viable vs. Non-Viable Myocardium. Pooled analysis of mean change in RWMS from eight studies comparing patients with viable and non-viable myocardium. The overall mean difference was –0.05 (95% CI: –0.45 to 0.35), indicating no statistically significant difference. Substantial heterogeneity was observed (I² = 100%, Tau² = 0.34). The result was notably influenced by a single outlier study reporting a large effect size. Abbreviations: SD – standard deviation; CI – confidence interval; IV – inverse variance; REML – restricted maximum-likelihood estimator.

### Sensitivity Analysis - RWMS Change

A separate pooled analysis was conducted using change in regional wall motion score (RWMS) as the outcome in eight studies [17–29]. The initial overall effect showed no significant difference between viable and non-viable myocardium, with a mean difference of -0.01 (95% CI: -0.51 to 0.49, p = 0.96), and marked heterogeneity (I² = 100%). However, sensitivity analyses revealed that this result was heavily influenced by a single outlier: the study by Murashita et al., [30], which reported an unusually large effect (mean difference 2.40 [95% CI: 1.32 to 3.48]). When this study was excluded, the pooled mean difference shifted to -0.27 (95% CI: -0.41 to -0.13, p = 0.0002), indicating a statistically significant improvement in RWMS among patients with viable myocardium. This exclusion also substantially reduced heterogeneity (Tau² dropped from 0.49 to 0.03, I² from 100% to 97%). Other exclusions did not materially alter the overall effect, and no additional single study exerted a similar level of influence on the pooled outcome. These findings suggest that the beneficial effect of myocardial viability on RWMS may be obscured when high-impact outliers are included, highlighting the importance of rigorous sensitivity testing in meta-analyses of heterogeneous data.

### All-Cause Mortality

A meta-analysis of nine studies evaluated the association between myocardial viability and all-cause mortality following revascularization [17,18,19,20,21,22,23,24,25]. The pooled hazard ratio demonstrated a significantly higher risk of mortality in patients without viable myocardium compared to those with viability, with an overall hazard ratio of 2.42 (95% CI: 2.15 to 2.74; p < 0.00001) (Figure *5*). This suggests that patients with viable myocardium have markedly better survival outcomes after revascularization. However, heterogeneity was substantial (I² = 96%, χ² = 179.04, p < 0.00001), reflecting variability across studies in both methodology and outcome reporting. Two studies-Pagano et al., 1998 [26] and Slart et al., 2011 [27]-exhibited extreme hazard ratios with unusually wide confidence intervals (e.g., HR > 1000), indicating potential data distortion or computational anomalies. Despite these outliers, the direction and statistical significance of the pooled effect remained robust, underscoring the prognostic value of myocardial viability in informing revascularization decisions.

**FIGURE 5:**
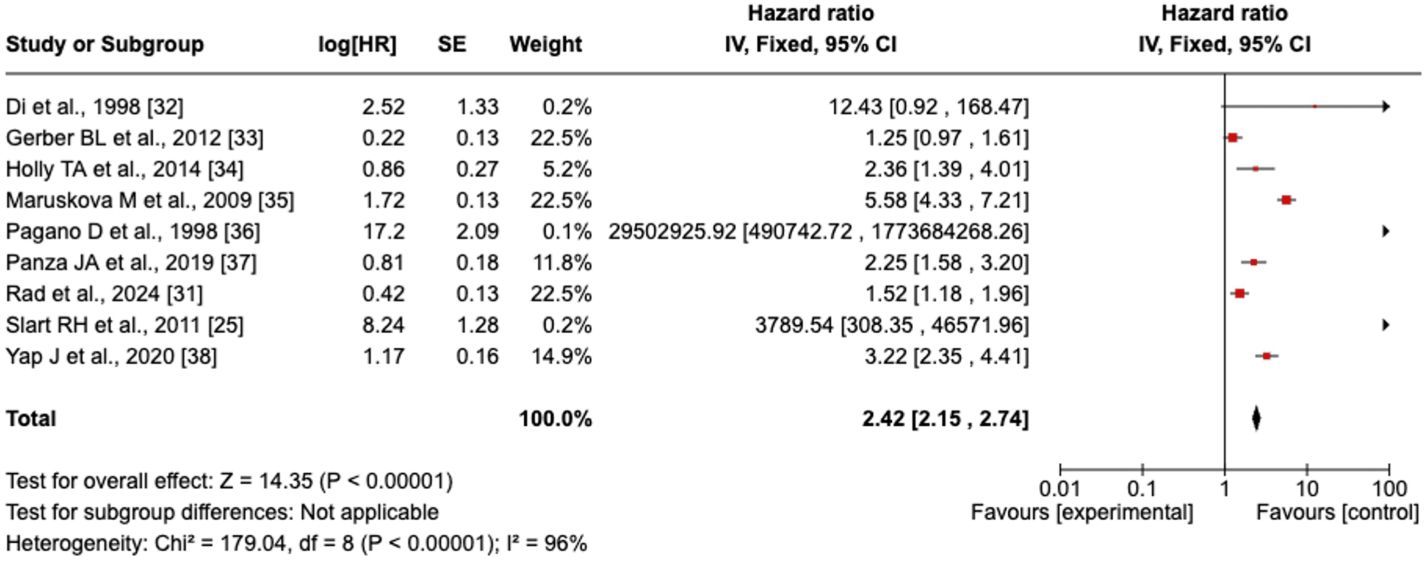
Forest Plot of All-Cause Mortality in Patients With Viable vs. Non-Viable Myocardium Following Revascularization. Pooled hazard ratios (HRs) from nine studies comparing all-cause mortality in patients with viable versus non-viable myocardium after revascularization. The overall HR was 2.42 (95% CI: 2.15–2.74), indicating significantly higher mortality in patients without viable myocardium. Two studies reported extreme HRs with wide confidence intervals, suggestive of computational anomalies or data skew. Despite heterogeneity (I² = 96%), the direction and statistical significance of the effect were consistent. Abbreviations: HR – hazard ratio; CI – confidence interval; IV – inverse variance; SE – standard error.

To assess the stability of this association, a leave-one-out sensitivity analysis was performed. Exclusion of individual studies yielded HRs ranging from 1.90 (95% CI: 1.66-2.18) to 2.94 (95% CI: 2.56-3.38), with all estimates remaining statistically significant. The strongest attenuation occurred when Maruskova et al., 2009 [28] was excluded (HR = 1.90), suggesting its high weight and effect size influenced the pooled result. Conversely, removing Yap et al., 2020 [29] produced the lowest pooled HR at 2.31 (95% CI: 2.02-2.63), indicating this study also contributed materially to the pooled effect size. Despite these fluctuations, the overall interpretation remained consistent-non-viable myocardium was robustly associated with increased mortality risk after revascularization. These findings emphasize the prognostic utility of viability imaging for long-term outcomes in ischemic cardiomyopathy.

### Risk of Bias Assessment

A total of 22 studies were included in the risk of bias evaluation-three randomized controlled trials [17–19] assessed using the RoB 2.0 tool and 19 observational studies [20–38] assessed using the ROBINS-I tool.

Among the randomized studies, two trials [17,18] were rated as having low risk of bias across all domains (Figure *6*). One trial [19] was judged to have some concerns due to limited clarity around the randomization process and potential bias in outcome measurement. No randomized trial was rated as having a high overall risk of bias.

**FIGURE 6:**
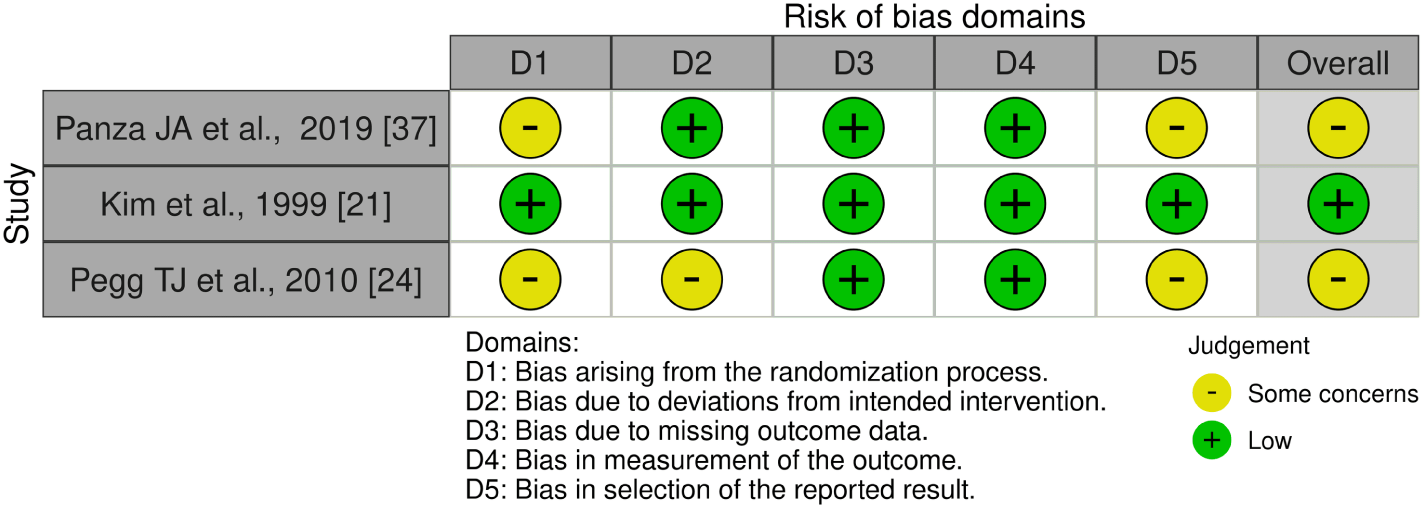
Risk of bias assessment using the ROB 2 tool for randomized studies. Traffic light plot summarizing the risk of bias across five domains for the three randomized controlled trials included in this review. Judgments are color-coded as low risk (green), some concerns (yellow), or high risk (red) based on the Cochrane ROB 2.0 tool. Most studies demonstrated low risk in domains related to outcome measurement and reporting, while concerns were most frequently noted in the randomization process and adherence to the intended intervention.

For the 19 observational studies, ROBINS-I assessments revealed that the majority (n = 14) [20–33] were judged to have moderate risk of bias, primarily due to concerns related to confounding and outcome measurement (Figure *7*). Five studies were considered to have a low risk of bias, while two studies were judged to be at serious risk of bias, driven by substantial baseline imbalances and incomplete adjustment for key prognostic factors.

**FIGURE 7:**
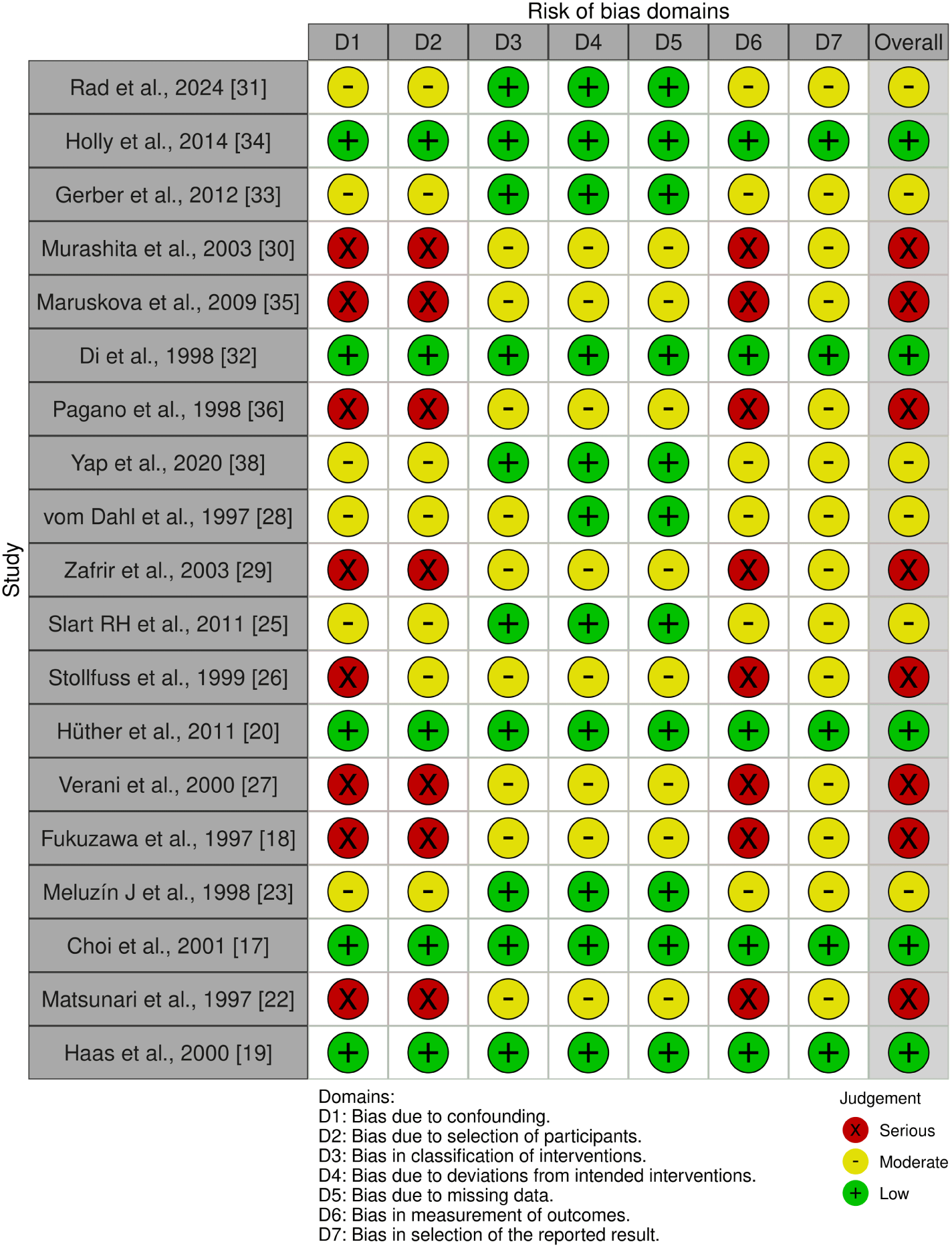
Risk of Bias Assessment Using the ROBINS-I Tool for Included Observational Studies. Traffic-light plot summarizing risk of bias across seven domains for observational studies included in the systematic review, assessed using the ROBINS-I tool. Each domain is color-coded: green (low risk), yellow (moderate risk), and red (serious risk). Domains evaluated include confounding, participant selection, intervention classification, deviations from intended interventions, missing data, outcome measurement, and reporting bias. Overall risk of bias was judged to be moderate in most studies, with serious risk identified in a minority due to confounding and selection bias. Abbreviations: ROBINS-I – Risk Of Bias In Non-randomized Studies of Interventions.

Overall, while a moderate level of bias was present in several observational studies, the directionality and consistency of findings across low-bias studies support the robustness of the pooled results (Figure *7*).

### Summary of Findings

This meta-analysis demonstrates that myocardial viability, as assessed by imaging, is significantly associated with improved outcomes in ischemic cardiomyopathy. Patients with viable myocardium showed greater improvement in LVEF following revascularization (mean difference 4.03%, 95% CI: 3.76-4.31), a finding that remained robust across imaging modalities and sensitivity analyses. The strongest improvements were observed with dobutamine echocardiography and CMR. Leave-one-out analyses confirmed that no single study altered the direction or significance of this association.

For regional wall motion score (RWMS), pooled analysis initially showed no significant difference between groups. However, sensitivity testing revealed that exclusion of one outlier (Murashita et al., 2003) yielded a significant benefit in RWMS for viable myocardium (mean difference -0.27, 95% CI: -0.41 to -0.13), suggesting results were obscured by heterogeneity.

In terms of prognosis, non-viable myocardium was associated with a significantly higher risk of all-cause mortality post-revascularization (HR = 2.42, 95% CI: 2.15-2.74). Sensitivity analysis confirmed this relationship remained stable (range: HR 1.90-2.94), with no exclusions reversing the overall effect.

Altogether, viability imaging consistently identifies patients more likely to benefit functionally and prognostically from revascularization, supporting its role in clinical decision-making.

## Discussion

This systematic review and meta-analysis synthesize the evidence on whether myocardial viability, assessed by advanced imaging, predicts improved outcomes following coronary revascularization. Our findings build on the foundational STICH trial, which questioned the utility of viability testing in guiding revascularization [8]. In contrast to STICH, this review focuses specifically on post-revascularization outcomes among patients stratified by viable versus non-viable myocardium and incorporates more recent data, including contemporary imaging modalities and updated clinical endpoints. Three primary outcomes were analysed: change in left ventricular ejection fraction (LVEF), regional wall motion score (RWMS), and all-cause mortality.

The overall pooled data confirm that the presence of viable myocardium is strongly associated with improvement in LVEF after revascularization. This benefit was consistent across modalities, with the most pronounced effects observed in patients assessed using dobutamine stress echocardiography and cardiac magnetic resonance (CMR) [39]. These findings support the pathophysiological rationale of myocardial hibernation: that chronically under perfused but viable myocardial segments can recover function following revascularization [3,32] Sensitivity analyses, including leave-one-out approaches, confirmed that this LVEF benefit persisted despite the exclusion of studies with large weight or neutral findings, underscoring the robustness of this association.

However, our analysis of RWMS yielded more nuanced results. Initial pooled estimates showed no significant difference in RWMS change between viable and non-viable groups. Yet, exclusion of one influential outlier Murashita et al., [30] revealed a statistically significant improvement in RWMS in viable myocardium, indicating that heterogeneity and outlier effects may obscure underlying associations. This further highlights the importance of standardized definitions and careful methodological control when evaluating regional myocardial recovery Allman et al., [40].

The prognostic implications of myocardial viability were also evident in our mortality analysis. Patients with non-viable myocardium had a significantly higher risk of all-cause mortality following revascularization (HR = 2.42). While heterogeneity was substantial, the directionality of the effect remained unchanged in all sensitivity analyses. These findings challenge the interpretation of prior studies such as REVIVED-BCIS2, which did not incorporate viability imaging and found no survival benefit from PCI in ischemic cardiomyopathy. Our data suggest that imaging-based viability assessment may identify a subgroup of patients more likely to benefit not only functionally but also prognostically from revascularization.

This conclusion is further supported by recent observational studies that directly compared image-guided versus non-image-guided revascularization. The study by Boehm et al. [41], which stratified CABG candidates using FDG-PET, demonstrated significantly better perioperative and long-term survival in the imaging-guided group. These findings reinforce that the diagnostic yield of viability imaging may lie not only in identifying any viability but in characterizing its extent and pattern [42].

Moreover, Syed et al., [43] demonstrated that dobutamine stress MRI alone was sufficient to detect both ischemia and viability, potentially streamlining work-up in institutions with MRI capabilities. This aligns with our subgroup findings where dobutamine stress echocardiography showed the highest mean LVEF improvement, raising the possibility of greater physiological responsiveness in these patients.

Taken together, our review indicates that viability imaging remains a valuable tool in the management of ischemic cardiomyopathy. From a health economics perspective, imaging could potentially be reserved for cases where revascularization is being considered but clinical equipoise exists. Identifying viable myocardium could justify intervention in high-risk patients, while confirming non-viability may help avoid futile procedures, reduce costs, and redirect care toward optimal medical therapy or advanced heart failure interventions (Di MF et al., and Beanlands et al., and Perera et al., [42,4,44].

Limitations of this review include the observational nature of most included studies, reliance on imputed or author-calculated data in some cases, and the absence of randomized trials directly comparing viability-guided versus angiography-guided decision-making. While we employed rigorous statistical methods and sensitivity analyses, residual confounding cannot be excluded. Furthermore, variation in imaging protocols, thresholds for viability (Ralota et al., [45], and timing of follow-up contribute to heterogeneity and limit direct comparison across studies.

## Conclusions

In this systematic review and meta-analysis, we demonstrate that myocardial viability assessment using advanced imaging modalities meaningfully stratifies patients with ischemic cardiomyopathy undergoing revascularization. Patients with viable myocardium experienced significant improvements in left ventricular ejection fraction (LVEF) and regional wall motion score (RWMS) and demonstrated a markedly lower risk of all-cause mortality compared to those without viability. These findings suggest that viability imaging holds substantial clinical value, helping to identify patients who are most likely to benefit from revascularization both functionally and prognostically. The consistency of results across imaging modalities, along with the robustness observed in multiple sensitivity analyses, supports the integration of viability testing into revascularization decision-making processes.

Given the substantial morbidity and mortality associated with ischemic cardiomyopathy, refining patient selection for revascularization is critical to improving outcomes. Viability imaging offers an opportunity to tailor treatment strategies to individual patient profiles, potentially avoiding futile interventions in patients with extensive non-viable myocardium while prioritizing revascularization in those most likely to derive meaningful benefit. In doing so, it reinforces a patient-centered approach that balances procedural risk against potential functional and survival gains. Future research should focus on standardizing imaging thresholds, exploring the comparative effectiveness of different imaging modalities, and conducting randomized trials to further refine the role of viability imaging in clinical practice. Overall, these results advocate for a selective but strategic use of viability assessment to optimize patient outcomes in ischemic cardiomyopathy.

## Appendices

### Search Strategy

**TABLE 1:**
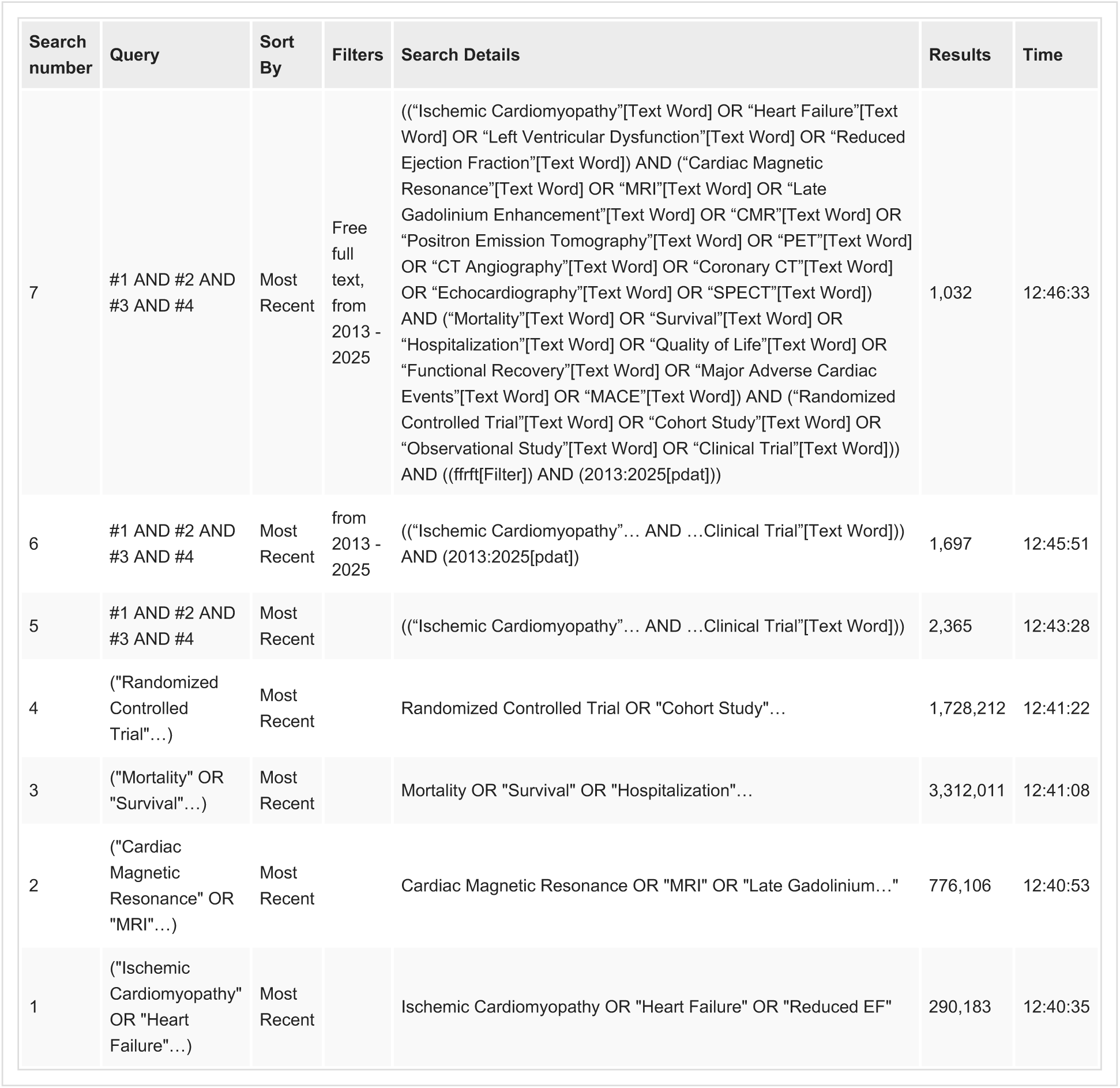
PubMed search strategy used for identifying relevant studies on myocardial viability imaging and coronary revascularization in ischemic cardiomyopathy. The table outlines sequential search steps combining terms related to ischemic cardiomyopathy, imaging modalities (e.g., CMR, PET, SPECT), clinical outcomes (e.g., mortality, quality of life), and study types (e.g., randomized controlled trials, cohort studies). Filters were applied for publication date (2013–2025) and free full-text availability, yielding a final result of 1,032 studies at Search #7.

**TABLE 2:** Embase search strategy used in the systematic review. This table outlines the sequential search strategy conducted in Embase (Ovid platform, 1974 to January 3, 2025) to identify studies evaluating outcomes of revascularization in patients with ischemic cardiomyopathy stratified by myocardial viability. Search terms included combinations of keywords related to cardiac dysfunction (e.g., ischemic cardiomyopathy, heart failure), imaging modalities (e.g., MRI, PET, SPECT), clinical outcomes (e.g., mortality, quality of life), and study design (e.g., randomized controlled trial, cohort). Boolean operators were applied to combine search sets, followed by limiting to full-text articles published from 2013 onwards. A total of 941 records were retrieved from Embase for screening.

| Search Number | Query | Results |
| --- | --- | --- |
| 1 | (ischemic cardiomyopathy or heart failure...) | 639,545 |
| 2 | (cardiac magnetic resonance imaging or MRI...) | 1,374,455 |
| 3 | (positron emission tomography or PET...) | 298,347 |
| 4 | (computed tomography angiography or CTA...) | 47,534 |
| 5 | (echocardiography or SPECT...) | 572,310 |
| 6 | (mortality or survival or hospital admission...) | 5,002,281 |
| 7 | (randomized controlled trial or cohort study...) | 3,264,061 |
| 8 | 2 or 3 or 4 or 5 | 2,092,638 |
| 9 | 1 and 6 and 7 | 64,423 |
| 10 | 8 and 9 | 8,975 |
| 11 | limit 10 to yr="2013 -Current" | 7,087 |
| 12 | limit 11 to full text | 941 |

## Data Availability

All data produced in the present work are contained within the manuscript and its supplementary materials. The data were extracted from publicly available published studies, all of which are cited and referenced.

## Acknowledgements

The authors would like to acknowledge Hasan Butt and Maria Taiwo for their substantial contributions to the development of this manuscript. Although authorship is limited to ten individuals due to journal restrictions, their involvement in the analysis and interpretation of data merits recognition, and they should be considered as contributing authors to this work.

